# Oral-motor complexity influences center of pressure patterns in adults with stroke-related communication disorders

**DOI:** 10.1101/2024.09.12.24313557

**Authors:** Daria Pressler, Sarah Dugan, Amu De Silva, Michael A. Riley, Sarah M. Schwab-Farrell

## Abstract

People with stroke (PwS) often exhibit altered postural control, and concomitant stroke-related communication disorders (e.g., aphasia, dysarthria) may be an underrecognized risk factor for post-stroke falls. This heightened fall risk may be related to alterations in postural control that emerge during different speaking and listening conditions. This study evaluated how variations in the relative articulatory demands during speech production—termed “oral-motor complexity”—affect postural center of pressure (COP) patterns among PwS, both with communication disorders (PwS-CDis) and without (PwS). Three groups of adults (PwS, PwS-CDis, and a nondisabled Control group) stood on a force platform while completing four 30-second quiet stance trials, followed by twelve 30-second trials randomized across three experimental conditions of varying oral-motor complexities (“ba”, “puh tuh kuh”, “rah shah lah nah”). COP variability (SD) was significantly higher during experimental conditions compared to quiet stance, regardless of group and movement plane. Differences in nonlinear time-dependent metrics were found across oral-motor task conditions, particularly among PwS-CDis, suggesting oral-motor complexity may influence underlying postural-motor organization. Distinct temporal-dynamical patterns observed in PwS-CDis indicate a possible link between pathology, postural control, and speech motor tasks, which is relevant when evaluating postural control in individuals with stroke-related communication disorders.

## INTRODUCTION

Individuals after stroke commonly demonstrate balance and postural control impairments and limitations, including altered center of pressure (COP) magnitude (de Haart et al., 2004), increased COP regularity (Schwab et al., 2023), and diminished anticipatory and reactive postural responses (Dickstein et al., 2004; Tasseel-Ponche et al., 2015). Specifically, people with stroke (PwS) show increased sway magnitude in the frontal and sagittal planes, which is more pronounced on the paretic side and during complex postural and cognitive tasks (Dickstein & Abulaffio, 2000; Mehdizadeh et al., 2015). PwS are generally at increased risk for falls, and fall risk is especially heightened for PwS with stroke-related communication disorders (e.g., dysarthria, aphasia) (Sullivan & Harding, 2019), indicating that communication disorders may be under-recognized as an important fall risk factor after stroke.

This heightened fall risk might be related to alterations in postural control known to exist when individuals (with and without disability) concurrently engage in tasks like speaking and listening. Even young adults exhibit increased sway frequency and amplitude during speech articulation tasks in contrast to a reduction in sway amplitude that occurs during dual-task (concurrent cognitive and postural) performance without concurrent articulation (Dault et al., 2003; Yardley et al., 1999). It may be that the increase in postural sway parameters when speaking derives from purely mechanical influences of speaking (e.g., movements of the jaw and head, altered respiratory-related trunk movements). However, speech-language processes also present specific neurocognitive demands such as those associated with planning and executing speech movements or with comprehending spoken language. As one example of the impact of such processes on balance, middle-aged adults with and without hearing impairments affecting speech recognition demonstrate increased COP excursion under challenging listening conditions when performing dual-tasks involving postural control and listening to speech (Helfer et al., 2020).

Similar dual-task findings involving concurrent postural control and speech have been observed in individuals with neurological conditions. For example, individuals with Parkinson’s disease and concomitant mild to moderate dysarthria (i.e., weakness of muscles used for speech) have been found to exhibit impaired motor and speech performance during concurrent speech articulation and postural-motor tasks (Dromey et al., 2010). Additionally, compared to an older adult control group, PwS without communication disorders exhibit greater postural control effects (e.g. increased path length) during the repetitive verbal utterance of “ba” than when sitting still, with no further increase in these effects during a more complex word generation task (e.g., verbally generating as many examples from a given category as possible within a specific time constraint) (Harley et al., 2006). These results suggest that performing a simple verbal utterance alone (“ba”) is sufficient to alter seated postural activity in PwS without communication disorders.

It is currently unknown how verbal utterance affects postural control among PwS who have stroke-related communication disorders—an especially important consideration given the potentially higher risk for falls in this population. Further, much of the prior dual-task research involving postural control and speaking in neurological disability has generally focused on verbal tasks with a cognitive component, such as lexical processing, word generation, and word retrieval tasks (e.g., Harley et al., 2006; Jehu et al., 2020). The impact of varying complexity levels of *non-cognitive* verbal utterances (e.g., “ba” vs. “puh tuh kuh”) on postural control after stroke, while importantly controlling for articulatory factors that could mechanically influence postural control sway measures (e.g., syllabic nucleus (“ah”) of utterances, number of phonemes in syllabic onset position), remains largely unknown.

In this paper, we categorize these verbal utterances based on their levels of “oral-motor complexity.” Oral-motor complexity refers to the relative articulatory complexity of the task dynamics—the physical and mechanical properties involved in the execution and completion of a specific task or action—involved in producing different syllables in a language, in this case, American English. Although complexity of task dynamics in speech can be described in different ways, in this study, we consider complexity levels as defined by the clinical definition of complexity: More complex syllables consist of speech sounds (like “r”) that lead individuals with articulatory deficits and communication disorders to make errors due to the number, location, and timing of articulatory movements required to make them (Campbell et al., 2010; Romani et al., 2017). These more complex speech sounds are also typically acquired later in childhood due to their complexity of articulation and are more likely to be part of a speech disorder, even across languages (Boyce, 2015). Understanding how speech motor movements of varying complexity impact postural control can provide important insights into the non-cognitive aspects of speech production (which are often difficult to disentangle from cognitive aspects) that may alter postural control after stroke, particularly among PwS who have concomitant communication disorders. Furthermore, exploring the connection between speech motor ability (revealed through different oral-motor tasks) and postural control may offer important opportunities for enhanced clinical collaboration after stroke, particularly between speech-language pathologists and physical therapists (Schwab, Dugan, & Riley, 2021).

The objective of this study was to evaluate the influence of verbal utterances with varying oral-motor complexities on the standing postural control (specifically COP patterns) of PwS, both with and without stroke-related communication disorders, compared to an age-matched control group without a history of stroke. We hypothesized that PwS with communication disorders would exhibit differences in COP dynamics (in both magnitude and temporal regularity) compared to PwS without such impairments and to age-matched controls, and that those differences would be more pronounced with increasing levels of oral-motor complexity.

## METHODS

### Participants

A sample of twenty individuals with a diagnosis of chronic stroke (stroke >6 mo prior to enrollment) scoring 0-3 on the Modified Rankin Scale (Rankin, 1957) were divided into two subgroups: Eleven individuals with stroke-related communication disorders or impairments (PwS-CDis) and nine individuals without communication disorders or impairments (PwS). Additionally, ten age-matched adults with no history of communication or neurological disorders were included as a control group (Control). Given that this study was first-of-its-kind, there was no sound basis for an a priori power analysis. The study procedure and recruitment method were reviewed and approved by the University of Cincinnati Institutional Review Board, and all participants provided written informed consent.

Participants in the PwS-CDis group were required to be fluent in English, to have a self-reported stroke-related communication disorder or impairment at time of enrollment (i.e., chronic communication disorder), and be able to maintain standing balance for ≥2 min with no physical support or assistive devices, except lower limb orthoses. Individuals were permitted to be active in physical therapy, occupational therapy, and/or speech therapy. Exclusion criteria included serious intellectual disability limiting the ability to follow simple one-step commands, deafness, blindness, severe motor impairments precluding experimental task performance, pain with weightbearing > 4/10 on the Numeric Pain Rating Scale (NPRS), pregnancy, seizure disorders, additional neurological conditions, or significant musculoskeletal conditions (e.g., amputation) which would preclude the valid administration of study measures. The PwS group met the same inclusion/exclusion criteria as the PwS-CDis group, but it did not include individuals with chronic communication disorders, and participants were not permitted to be enrolled in speech therapy. Importantly, this ensured functional comparability with the PwS-CDis group in all ways, except in communication abilities. The Control group met the same criteria as the PwS group but additionally did not have a history of neurological conditions. See Table 1 for complete demographics.

**Table 1.** Participant Characteristics. SD: Standard Deviation; PROMIS: Patient-Reported Outcomes Measurement Information System by NIH to assess physical, mental, and social health; PROMIS Mobility: Subset of PROMIS, score range 11-55, higher score = lower disability; PROMIS Speech: Subset of PROMIS, score range 7-35, higher score = higher disability.

|  | PwS (*n* = 9) | PwS-CDis (*n* = 11) | Control (*n* = 10) |
| --- | --- | --- | --- |
| Age, y (Mean ± SD) | 61 ± 8.3 | 61.5 ± 8.1 | 64.2 ± 8.8 |
| Gender, n (%) |  |  |  |
| Female | 4 (44.4) | 8 (72.7) | 5 (50.0) |
| Male | 5 (55.6) | 3 (27.3) | 5 (50.0) |
| Race, n (%) |  |  |  |
| African American/Black | 3 (33.3) | 4 (36.4) | 1 (10.0) |
| Asian/Pacific Islander | 0 (0.0) | 0 (0.0) | 1 (10.0) |
| Caucasian | 6 (66.7) | 7 (63.6) | 8 (80.0) |
| Height, m (Mean ± SD) | 1.7 ± 0.12 | 1.7 ± 0.10 | 1.7 ± 0.11 |
| Weight, kg (Mean ± SD) | 88.3 ± 16.8 | 80.8 ± 19.5 | 87.0 ± 17.8 |
| Stroke Age, mo. (Mean ± SD) | 91.4 ± 31.5 | 83.5 ± 71.7 |  |
| Stroke Type, n (%) |  |  |  |
| Ischemic | 5 (55.6) | 8 (72.7) |  |
| Hemorrhagic | 3 (33.3) | 2 (18.2) |  |
| Ischemic + Hemorrhagic | 0 (0.0) | 1 (9.1) |  |
| Unknown | 1 (11.1) | 0 (0.0) |  |
| Paretic Side, n (%) |  |  |  |
| Left | 3 | 0 (0.0) |  |
| Right | 4 | 10 (90.9) |  |
| Unknown/None | 2 | 1 Both |  |
| Modified Rankin Score, n (%) |  |  |  |
| 1 (No significant disability) | 4 (44.4) | 3 (27.3) |  |
| 2 (Slight disability) | 5 (55.6) | 3 (27.3) |  |
| 3 (Moderate disability) | 0 (0.0) | 5 (45.5) |  |
| PROMIS Mobility | 51.2 ± 3.87 | 46.27 ± 7.98 | 54.3 ± 2.21 |
| PROMIS Speech | 7.56 ± 1.42 | 16.55 ± 5.63 | 6.5 ± 0.7 |
| Hours/Week in Physical Occupational or Speech Therapy, n (%) |  |  |  |
| No therapy | 7 (77.8) | 7 (63.6) | 9 (90.0) |
| < 1 hour | 1 (11.1) | 0 (0.0) | 1 (10.0) |
| 1-2 hours | 1 (11.1) | 1 (9.1) | 0 (0.0) |
| 2-3 hours | 0 (0.0) | 2 (18.2) | 0 (0.0) |
| 3-4 hours | 0 (0.0) | 1 (9.1) | 0 (0.0) |
| Highest Level of Education, n (%) |  |  |  |
| High school diploma | 3 (33.3) | 0 (0.0) | 1 (10.0) |
| Some college | 4 (44.4) | 3 (27.3) | 1 (10.0) |
| Associate degree | 1 (11.1) | 0 (0.0) | 0 (0.0) |
| Bachelor's degree | 1 (11.1) | 3 (27.3) | 4 (40.0) |
| Graduate degree | 0 (0.0) | 5 (45.5) | 4 (40.0) |
| Median Annual Household Income, n (%) |  |  |  |
| < \$20,000 | 3 (33.3) | 2 (18.2) | 1 (10.0) |
| \$20,000-40,000 | 0 (0.0) | 1 (9.1) | 0 (0.0) |
| \$40,000-\$60,000 | 1 (11.1) | 0 (0.0) | 0 (0.0) |
| \$60,000-\$80,000 | 2 (22.2) | 2 (18.2) | 3 (30.0) |
| \$80,000-\$100,000 | 0 (0.0) | 1 (9.1) | 1 (10.0) |
| > \$100,000 | 0 (0.0) | 2 (18.2) | 4 (40.0) |
| Prefer not to say | 3 (33.3) | 3 (27.3) | 1 (10.0) |
| Other medical conditions (n) | Diabetes mellitus (3); Visual Impairment (1); Hyperlipedemia (1) | Diabetes mellitus (2); Hypertension (3); Heart Condition (2); Hearing loss (1) | Diabetes mellitus (1); Hypertension (1); Kidney Disease (1); Orthopedic condition (1) |

### Materials and Methods

Participants, while wearing comfortable footwear, stood on a force platform (AMTI Optimized AccuSway; Advanced Mechanical Technology, Watertown, MA) in a comfortable stance with feet approximately shoulder-width apart. AMTI *Balance Clinic* software recorded the COP in the *x* (medio-lateral) and *y* (anterior-posterior) directions at a sampling frequency of 100 Hz. All participants (including those in the Control group) used a fall-prevention overhead harness (Biodex Medical Systems, Shirley, NY). The harness provided no physical/body-weight support; it was implemented only for participant safety. During all sessions audio was recorded via computer integrated audio/video (Windows 12 Camera; Microsoft Corp., Redmond, WA).

While standing on the force platform, participants were instructed to look at an anteriorly fixed point in their line of sight with arms resting comfortably at their sides. Participants first completed four 30 s quiet stance trials to establish baseline measurements of postural control without experimental manipulation, followed by the experimental task. The experimental task consisted of three repetitive verbal utterance conditions (four 30 s trials of each) with varying levels of oral-motor complexity (additional details can be found in the next section): “ba,” “puh tuh kuh,” and “rah shah lah nah.” Experimental trial order was randomized. Participants were instructed, given individual impairments, to perform repetitive verbal utterances at a comfortable yet consistent rate with regular breathing to minimize substantial breaks in the sequence. Prior to the study start, the entire task was explained, and participants practiced each utterance sequence with prompts to maintain a consistent rate within and across conditions. This practice also ensured that the task novelty would not impact results and that participants could accurately reproduce each sequence.

Prior to each trial, participants were again prompted with the appropriate utterance and allowed to practice. Throughout all trials, a sign displaying the verbal utterances was posted at eye height on the wall, ensuring participants could clearly see the utterances, with confirmation obtained prior to starting the trials. The design aimed to minimize cognitive load and ensure participants attempted the correct verbal utterance sequence. In the PwS-CDis group, COP was recorded even in instances where an individual’s communication disorder precluded clear articulation/pronunciation of the verbal utterance sequence.

### Verbal Utterances & Complexity

The complexity levels of oral-motor tasks are commonly defined using the clinical concept of complexity, where speech sounds (phonemes) that are late-acquired or commonly misarticulated are considered to be more complex than others (Boyce, 2015; Romani et al., 2017; Shriberg et al., 1994). Another dimension of complexity is *syllabic complexity*, where syllables are considered more complex if they have a greater number of phonemes in the syllable (e.g. “street” is more complex than “tee”). In this study, syllabic complexity was held constant, in that each target utterance had the same syllabic nucleus (“ah”) and the same number of phonemes in syllabic onset position (one phoneme in onset, e.g., the “b” in “ba”). In addition, all syllables were permissible in American English, and included phonemes characteristic of American English (the language of all participants).

The complexity of the syllable targets varied along two dimensions: The number of different syllables in a sequence and the articulatory complexity of the consonants in the syllables. All sequences were semantically nonsensical to avoid possible cognitive processing influences on postural control (Davie et al., 2012). The oral-motor tasks we included in this study are described as follows:

- **Level One (Easiest, lowest complexity): “ba”** - This level involves a single, repeated syllable with minimal tongue movement. It is characteristic of early infant babbling due to its ease of production.
- **Level Two (Harder, medium complexity): “puh tuh kuh”** - This level involves articulation of three distinct syllables, each requiring unique articulatory movements: lip closure (“puh”), tongue blade/tip closure against the alveolar ridge (“tuh”), and tongue dorsum closure along the palate (“kuh”). These early-developing sounds in children are produced by complete occlusion and release of airflow in the vocal tract. The sequence is used in diadochokinetic tests for oral-motor control and correlates with speech disorder severity (Samlan & Weismer, 1995; Prathanee et al., 2003). Notably, individuals with balance disorders have been found to have different diadochokinetic rates than individuals without such balance disorders (Ackermann et al., 1995; Duffy, 2013).
- **Level Three (Hardest, greatest complexity): “rah shah lah nah”** - This level involves four different syllables, including two sonorant continuants with multiple vocal tract constrictions (“r” and “l”) and one fricative continuant (“sh”)^1^ which are all part of the later-acquired “late 8” consonants in typically developing children, and one nasal continuant (“n”) (Shriberg et al., 1994). Articulation at this level requires more finely-timed tongue movements and velum adjustments, imposing greater demands on the motor control system due to its inherent complexity.

### Outcome Measures

Custom R Scripts (version 4.3.1) were utilized for data processing and analysis. Artifacts in COP regularity metrics can be introduced by using data or signal processing techniques such as filtering or down-sampling (Rhea et al., 2015; Lubetzky et al., 2018). Therefore, we used unfiltered COP data in our final analysis to avoid such issues.

We calculated the standard deviation (SD) of the COP (cm) separately for the medio-lateral (ML) and anterior-posterior (AP) directions (*x* and *y* directions, respectively) as a standard, time-independent metric of the variability of COP trajectories.

We further subjected the COP data to two nonlinear time-series analysis methods, Recurrence Quantification Analysis (RQA) and Sample Entropy (SampEn), to characterize the regularity, predictability, and adaptability of the postural control system as reflected in COP dynamical patterns. While time-independent metrics like the SD of the COP are informative for determining differences in the amount/magnitude of sway variability, nonlinear time-series metrics determine if, in addition to (or instead of), differences exist in the temporal dynamics of postural sway (i.e., how the COP is varying over time, rather than just how much the COP varied). A number of studies have recommended against exclusively indexing postural variability with time-independent metrics without consideration of the dynamic patterns of the COP (Newell et al., 1993; Riccio, 1993; Riley, 2001; Riley & Turvey, 2002; Slifkin & Newell, 1998; van Emmerik & van Wegen, 2000, 2002), and some postural control studies have found differences in COP dynamics that are not apparent in traditional, time-independent variability measures (e.g., Schmit et al., 2005). This is a relevant consideration for the purpose of the present study as temporal patterns in motor behavior are reflective of the underlying control and organization of the perceptual-motor system (e.g., Newell et al., 1993; Riley et al., 2011; Riley, 2001; Riley & Turvey, 2002; van Emmerik & van Wegen, 2000). Thus, differences found in the temporal patterns of sway across oral-motor tasks or between groups would suggest differences in the underlying organization of postural control.

RQA (Webber & Zbilut, 2005) was used to assess COP stability and regularity. RQA quantifies the patterning, non-stationarity, and complexity of time series through the analysis of local recurrences (i.e., repetitions of states) in a reconstructed phase space (Riley et al., 1999). Recurrence plots (RPs) provide a visualization of patterns of recurrence, where a single point on the RP indicates a recurrence (a value has repeated itself) and a line indicates continuing recurrence where a series of values were repeated (Coco & Dale, 2014). A variety of RQA metrics can be used to quantify these kinds of patterns. In this study, we specifically evaluated maximum line length (MAXLINE) and RQA entropy (RQAEn):

- **MAXLINE** is the length of the longest diagonal line segment on the RP, indicating the maximum duration of time that the system is revisiting an area of reconstructed phase space, which reflects the maximum length of a series of repeated data values. Higher MAXLINE indicates a higher degree of *mathematical* stability in a system’s behavior over time; mathematical stability refers to a dynamical system’s response to a slight change in initial conditions or to a trajectory perturbation and is distinct from (and should not be confused with) postural stability (Schmit et al., 2005). Prior research in Parkinson’s disease indicates that higher values of MAXLINE (compared to control participants) are characteristic of balance impairments and an overall tendency for increased regularity and rigidity of movement (Schmit et al., 2006).
- **RQAEn** was used as a measure of the irregularity of the deterministic structure of the COP data. Higher RQAEn values indicate that the distribution of the lengths of diagonal lines (or the lengths of time periods of repeated postural sway) has become less predictable, or that there are more diagonal lines of different lengths (Haddad et al., 2008). In other words, the postural system repeats many different subsets of the length of its behavior (i.e., more “structured variation” across behavioral modes) over time (Bonnette et al., 2020; Negahban et al., 2013). RQAEn should be carefully interpreted as it is not a *direct* quantification of the complexity of an analyzed time series and its behavior can be counterintuitive (Ramdani et al., 2013). Previous literature has found higher RQAEn in the ML direction for “fallers” compared to “non-fallers” (Ramdani et al., 2013). Further, individuals with multiple sclerosis and controls exhibit lower RQAEn when faced with greater postural or cognitive task demands (Negahban et al., 2013).

The R package ‘crqa’ (v.2.0.3) was used for calculating RQA metrics (Coco et al., 2021). RQA parameters were estimated for each individual trial using the routine of Coco and Dale (2014). Delay values ranged from 25 to 141 samples (*M* = 56.60, *SD* = 15.31), and embedding dimension ranged from 3 to 6 (*M* = 4.08, *SD* = 0.42). Radius was adjusted per trial to achieve a fixed recurrence rate between 2.25% and 2.5%, following recommendations of Wallot (2017) and Webber and Zbilut (2005). RQA metrics were also estimated for a subset of data using a fixed recurrence rate between 1.0% and 1.25% to ensure that the pattern of results was consistent across recurrence rates. The relative pattern of results was comparable across recurrence rates, confirming that results were not an artifact of parameter selection. Prior to phase space reconstruction and RQA calculations, signals were centered about their respective means. RQA metrics were calculated for the AP and ML directions separately.

SampEn complements RQA by measuring the likelihood that closely aligned sequences of data points will maintain their similarity in future comparisons over a set time interval. Lower SampEn values indicate more regularity and predictability, while higher values indicate more irregularity and unpredictability, providing important insights into the time-dependent structure of the COP. SampEn is distinct from RQAEn, and similarities between the two may be limited to semantics. For example, there is evidence that increases in SampEn can be observed with decreases in RQAEn (Rhea et al., 2011). It is important to recognize that RQAEn is an entropy algorithm applied to the distribution of diagonal line segments in the RP (as previously described), rather than to the time series directly (cf. SampEn). While RQAEn is similar to SampEn in that it measures the variability of repeated patterns in the time series, RQAEn is unique and provides non-redundant information from SampEn in that it measures the irregularity of the distribution of diagonal line lengths on the RP, reflecting the variability in how repeated patterns persist and evolve over time (Fino et al., 2016).

SampEn is formally calculated as the negative natural logarithm of the conditional probability that a vector similar to (within a tolerance, *r)* a template of length *m* will be found at length *m + 1*. The R package ‘TSEntropies’ (v.0.9) was used to estimate SampEn (Tomčala, 2020). Following recommendations of Montesinos et al. (2018) and Lubestzky et al. (2018) for COP data, SampEn was estimated using multiple parameter combinations to ensure that the pattern of results was consistent across parameter choices. Final SampEn values were determined with *m* = 3 and *r* = 0.1, and were additionally calculated at *m* = {2, 4} and *r* = {0.1, 0.3} yielding the same general pattern of results over those ranges. SampEn was calculated separately for the AP and ML directions.

### Statistical Analysis

Data were subjected to outlier analysis (± 2 MAD) prior to statistical analysis. For each outcome measure we employed mixed-effects modeling to assess the effect of group (PwS-CDis, PwS, Control), direction (AP, ML), and condition (quiet stance, “ba,” “puh tuh kuh,” “rah shah lah nah”) on COP patterns, considering both main effects and their interactions. In these models, group, direction, condition, and their interactions were treated as fixed effects, and participant was entered as a random effect. Model building followed a backward stepwise approach, evaluating model fit by comparing the deviance (−2 Log Likelihood; -2LL) between a larger model and a simpler nested model that excluded the predictor under analysis at each step, using the ‘step’ function within the R package ‘lmerTest’ (Kuznetsova et al., 2017). The change in -2LL follows a chi-square distribution with degrees of freedom (DOF) equal to the difference in the number of parameters between nested models, allowing a test of statistical significance. The final model retained only those main effects and interactions that significantly enhanced model fit. Significant interactions were further examined through simple-effects analysis, with DOF adjusted using Satterthwaite’s method.

## RESULTS

### COP Variability: SD of COP

There were significant main effects of condition, *F*(3, 860.93) = 23.31, *p* < .001, and direction, *F*(1, 862.01) = 970.29, *p* < .001. The SD of COP was higher in all experimental conditions compared to quiet stance, regardless of group and direction. SD of COP was higher in the AP direction (*M* = 0.43 cm, *SE* = 0.02 cm) compared to the ML direction (*M* = 0.21 cm, *SE* = 0.02 cm), regardless of group and condition. These main effects were qualified by a group × direction × condition interaction, *F*(6, 860.81) = 2.18, *p* < .001.

Because our research question and hypothesis were focused on understanding differences between conditions and groups, we sought to understand this three-way interaction by parsing by direction. In the AP direction, there was only a main effect of condition, *F*(3, 395.57) = 12.41, *p* < .001, such that SD of COP was higher in all experimental conditions compared to quiet stance (Figure 1).

**Figure 1.**
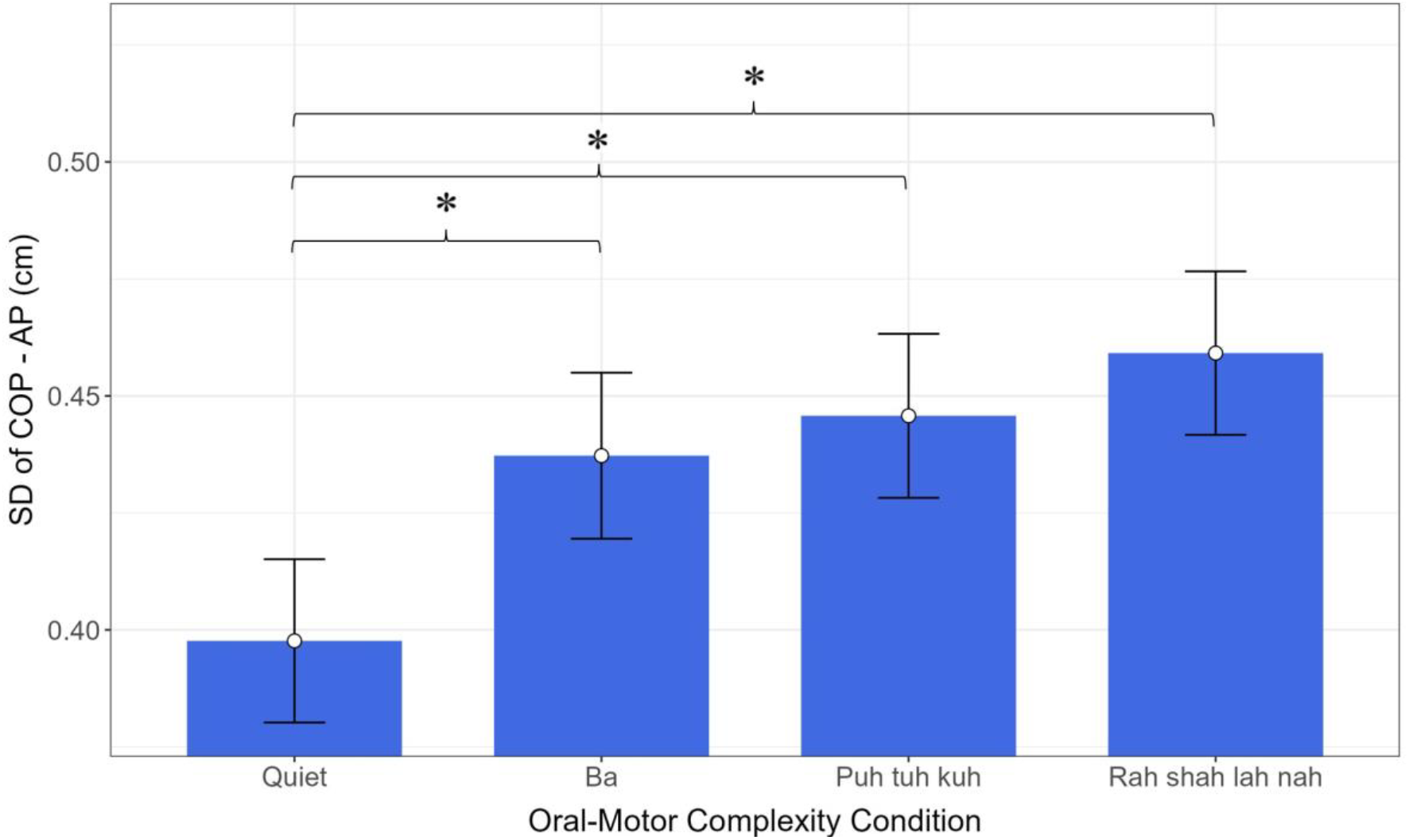
Anterior-posterior (AP) SD of COP by oral-motor complexity condition. Error bars represent the standard error of the mean (SEM). Asterisks (*) denote statistical significance (*p* < .05) between all experimental conditions and quiet stance.

In the ML direction, there was a main effect of condition, *F*(3, 435.12) = 15.79, *p* < .001, such that SD of COP was again higher in all experimental conditions compared to quiet stance. The main effect of condition in the ML direction was qualified by a group × condition interaction, *F*(6, 435.11) = 3.17, *p* < .001. When parsed by group, simple-effects analysis revealed differences between conditions in both the PwS and PwS-CDis groups, with higher ML SD of COP for all experimental conditions compared to quiet stance; the Control group did not demonstrate differences in ML SD of COP between conditions. When parsed by condition, simple-effects analysis revealed higher ML SD of COP in the PwS group compared to the Control group in both the “ba” and “rah shah lah nah” conditions. (Figure 2).

**Figure 2.**
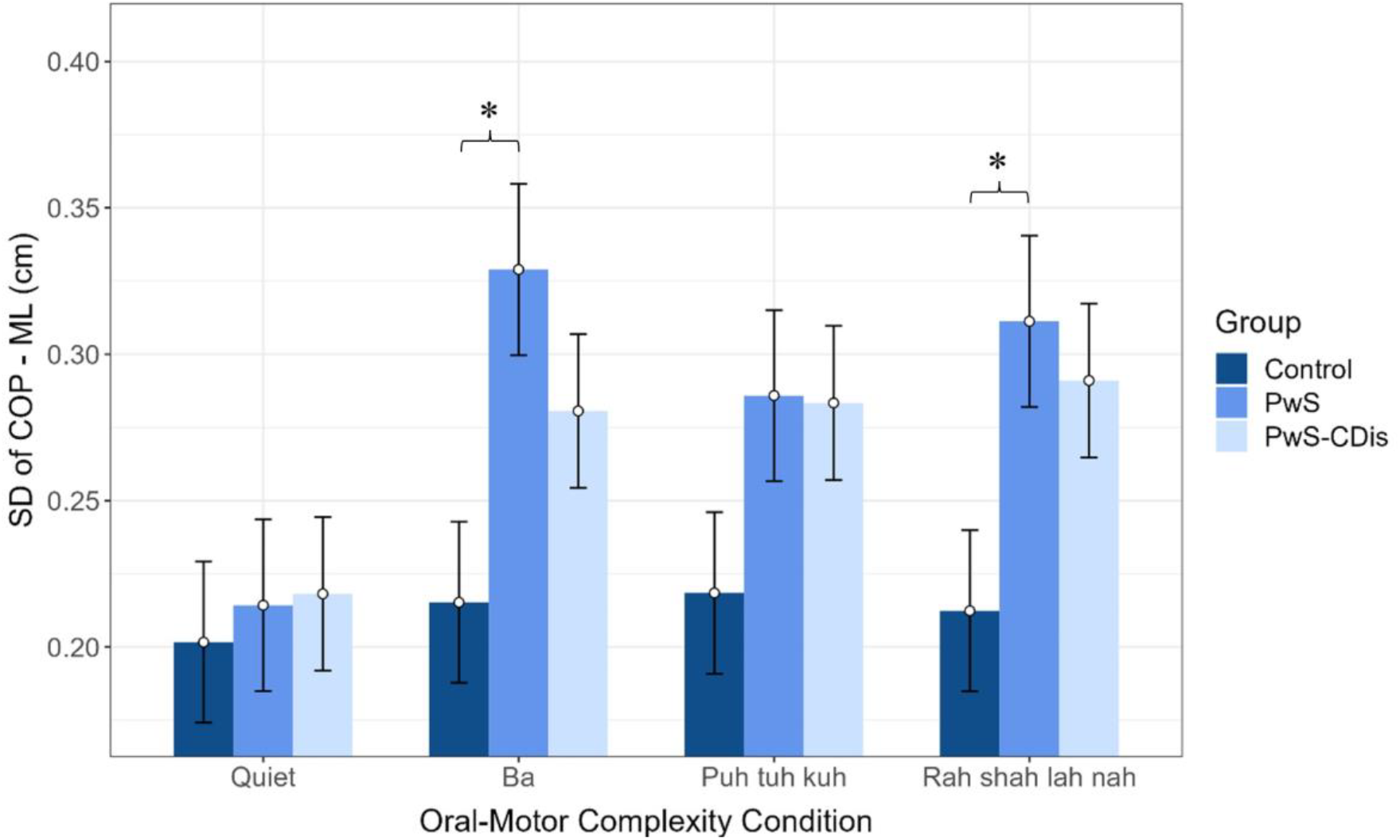
Medio-lateral (ML) SD of COP by oral-motor complexity condition for each group. Error bars represent the standard error of the mean (SEM). Asterisks (*) denote statistical significance (*p* < .05) between PwS and Control group for the “ba” condition and “rah shah lah nah” condition. The significant difference between all experimental conditions compared to quiet stance for the PwS and PwS-CDis groups is not indicated (see text for details).

Thus, the group × direction × condition interaction can be explained by increased AP SD of COP in all experimental conditions, regardless of group, and higher ML SD of COP in experimental conditions in the PwS and PwS-CDis groups (compared to quiet stance). Additionally, the PwS group exhibited greater ML SD of COP compared to the Control group specifically during the “ba” and “rah shah lah nah” conditions. The PwS-CDis group did not show statistical differences in SD of COP in either the ML or AP directions compared to the Control and PwS groups; rather, the PwS-CDis group only exhibited increased SD of COP (compared to quiet stance) in the ML and AP directions during experimental conditions.

### Mathematical Stability: MAXLINE

There were significant main effects of condition, *F*(3, 842.69) = 3.14, *p* = .02, and direction, *F*(1, 843.21) = 6.30, *p* = .01. MAXLINE was higher in the “ba” condition (*M* = 2810, *SE* = 3.92) compared to the “rah shah lah nah” condition (*M* = 2801, *SE* = 3.87), regardless of group or direction. MAXLINE was higher in the AP direction (*M* = 2809, *SE* = 3.53) compared to the ML direction (*M* = 2804, *SE* = 3.55), regardless of group or condition. These main effects were qualified by a group × condition interaction, *F*(6, 842.69) = 2.91, *p* = .008, and a group × direction interaction, *F*(2, 843.19) = 8.87, *p* < .001.

To understand the group × condition interaction, we parsed by both group and condition in separate simple-effects analyses. When parsed by group, simple-effects analysis revealed differences between conditions in the PwS-CDis group, such that MAXLINE was higher in the “ba” condition (*M* = 2823, *SE* = 6.32) compared to the “puh tuh kuh” condition (*M* = 2806, *SE* = 6.26) and the “rah shah lah nah” condition (*M* = 2805, *SE* = 6.21). When parsed by condition, there were differences between groups in quiet stance, such that MAXLINE was higher in the PwS group (*M* = 2819, *SE* = 6.11) and in the PwS-CDis group (*M* =2817, *SE* = 5.40) compared to the Control group (*M* = 2791, *SE* = 5.61). Additionally, MAXLINE in the “ba” condition was higher in the PwS-CDis group (*M* = 2823, *SE* = 6.08) compared to the Control group (*M* = 2796, *SE* = 6.32). Thus, the group × condition interaction is largely explained by the PwS-CDis showing a different pattern of results across experimental conditions in terms of mathematical stability: PwS-CDis showed more mathematical stability (i.e., less flexibility) in the “ba” condition compared to the other experimental conditions and compared to the Control group. The PwS and Control groups did not demonstrate changes in mathematical stability across experimental conditions (Figure 3).

**Figure 3.**
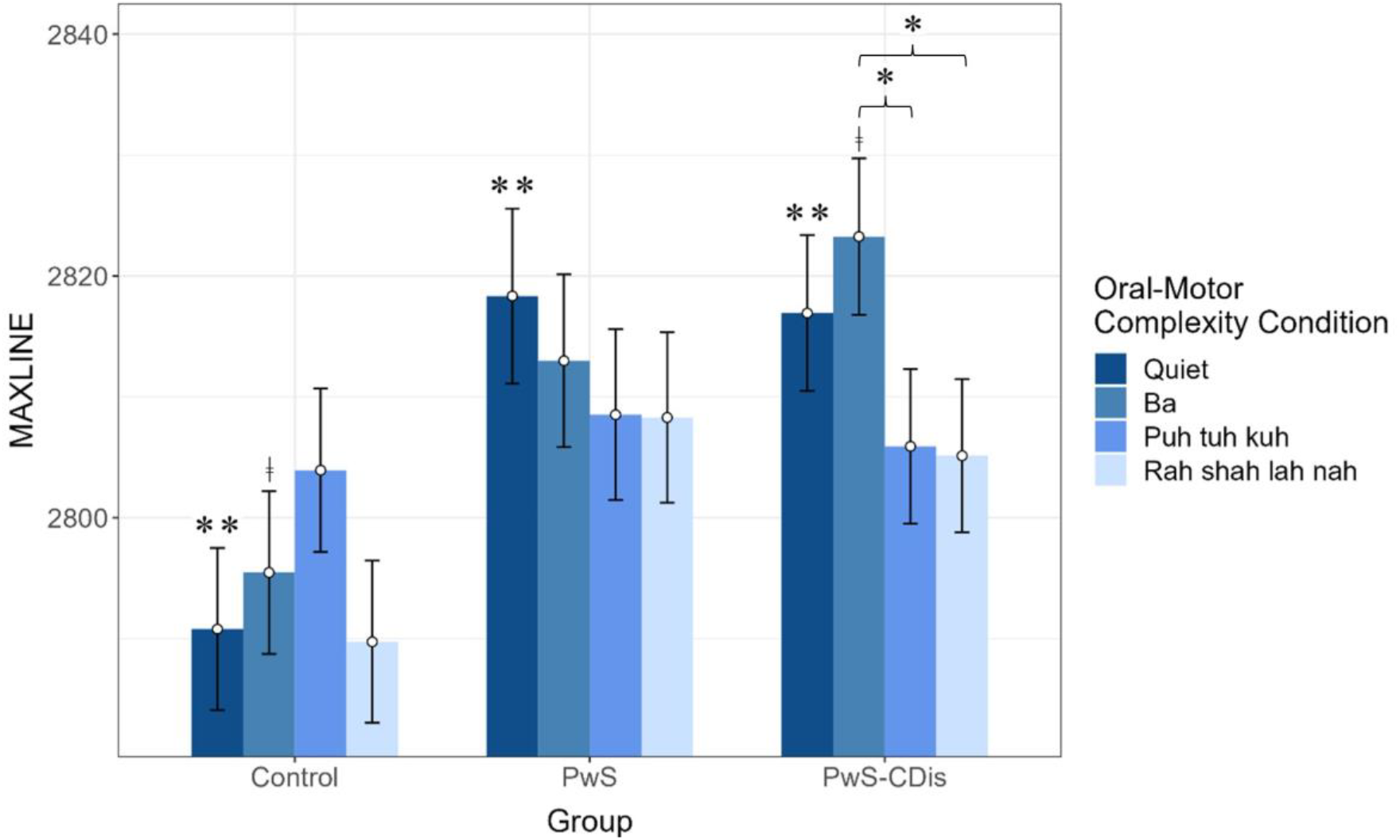
COP MAXLINE by group for each oral-motor complexity condition. Error bars represent the standard error of the mean (SEM). Statistical significance is denoted as *p* < .05. Asterisk (*) indicates significant difference between the “ba” condition compared to the “puh tuh kuh” and “rah shah lah nah” conditions in the PwS-CDis group. Double asterisks (**) indicate significant difference between the PwS and PwS-CDis groups compared to the Control group in quiet stance. Obelus (ǂ) denotes significant difference between the PwS-CDis group compared to the Control group in the “ba” condition.

To understand the group × direction interaction, we first parsed by group. MAXLINE in the Control group was significantly higher in the AP direction (*M* = 2804, *SE* = 5.92) compared to the ML direction (*M* = 2786, *SE* = 5.99). When parsed by direction, there were significant differences in MAXLINE between groups in the ML direction, such that MAXLINE was higher in the PwS-CDis group (*M* = 2815, *SE* = 7.21) compared to the Control group (*M* = 2785, *SE* = 7.58). These group × direction interaction results again suggest differences in mathematical stability found in the PwS-CDis group (Figure 4).

**Figure 4.**
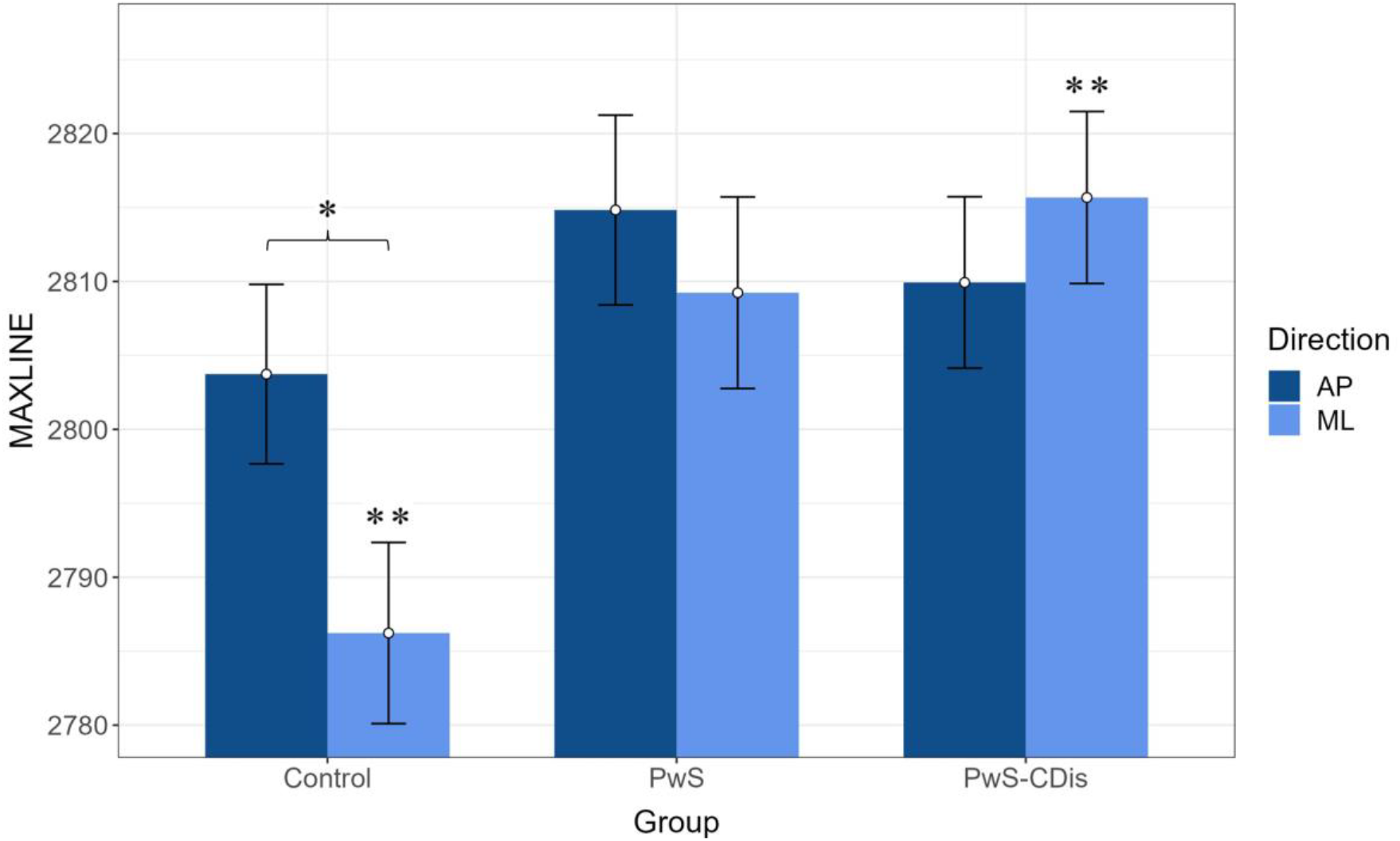
COP MAXLINE by group for AP and ML directions. Error bars represent the standard error of the mean (SEM). Statistical significance is denoted as *p* < .05. Asterisk (*) indicates significant difference between AP and ML in the Control group and double asterisks (**) indicate significant difference between the PwS-CDis group and Control group in ML direction.

### Structural Variation in Behavioral Modes: RQAEn

There were no main effects of group, condition, or direction; however, there was a significant group × condition interaction, *F*(6, 867.06) = 2.66, *p* = .01. When parsed by condition, simple-effects analysis revealed no differences between groups. However, when parsed by group, there were differences found between conditions in the PwS-CDis group, such that RQAEn was lower in the “ba” condition (*M* = 3.19, *SE* = 0.08) compared to quiet stance (*M* = 3.31, *SE* = 0.08) and the “rah shah lah nah” condition (*M* = 3.30, *SE* = 0.08). Thus, the group × condition interaction can be explained by the PwS-CDis group exhibiting more regular deterministic patterning when performing the “ba” condition compared to quiet stance and the “rah shah lah nah” condition (Figure 5).

**Figure 5.**
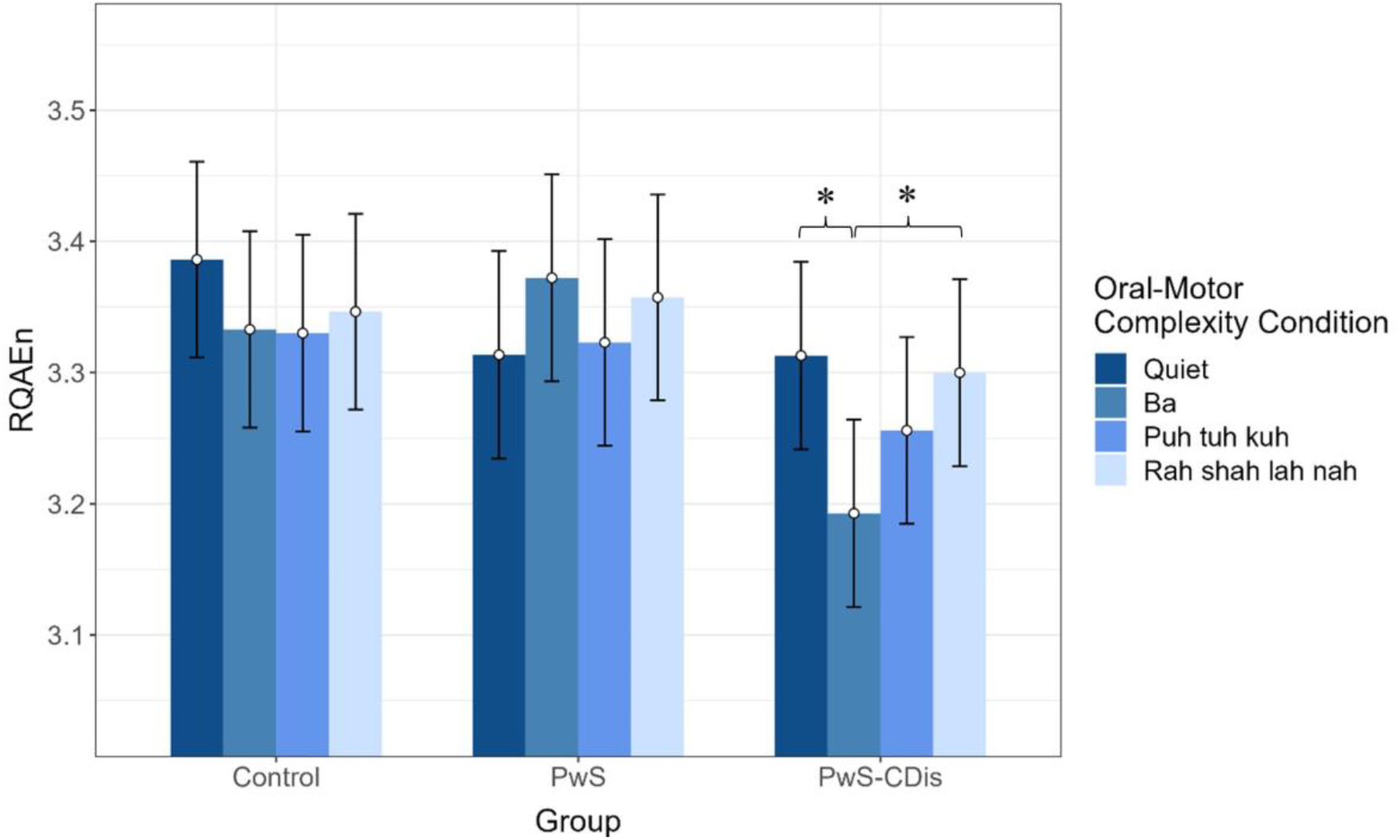
COP RQAEn by group for each oral-motor complexity condition. Error bars represent the standard error of the mean (SEM). Asterisks (*) denote statistical significance (*p* < .05) between the “ba” condition compared to quiet stance and the “rah shah lah nah” condition in the PwS-CDis group.

### Regularity & Predictability: SampEn

There was a significant main effect of condition, *F*(3,853.41) = 4.016, *p* = .007, such that SampEn was lower in the “rah shah lah nah” condition (*M* = 0.24, *SE* = 0.01) compared to the “puh tuh kuh” condition (*M* = 0.26, *SE* = 0.01) and quiet stance (*M* = 0.26, *SE* = 0.01). This main effect was qualified by a group × condition interaction, *F*(6, 853.41) = 2.77, *p* = .01.

When parsed by condition, simple-effects analysis revealed no differences between groups. However, when parsed by group, simple effects analysis showed differences between conditions in the PwS-CDis group, such that SampEn was significantly lower (i.e., more regular/predictable) in the “rah shah lah nah” condition (*M* = 0.26, *SE* = 0.02), compared to the “ba” condition (*M* = 0.29, *SE* = 0.02); the Control group also showed differences between conditions, such that SampEn was significantly lower in the “rah shah lah nah” condition (*M* = 0.21, *SE* = 0.02) compared to quiet stance (*M* = 0.25, *SE* = 0.02). Accordingly, the significant group × condition interaction can be explained by the PwS-CDis group showing a different pattern of SampEn results across conditions, while the Control and PwS group showed similar patterns of SampEn results across oral-motor task conditions (Figure 6).

**Figure 6.**
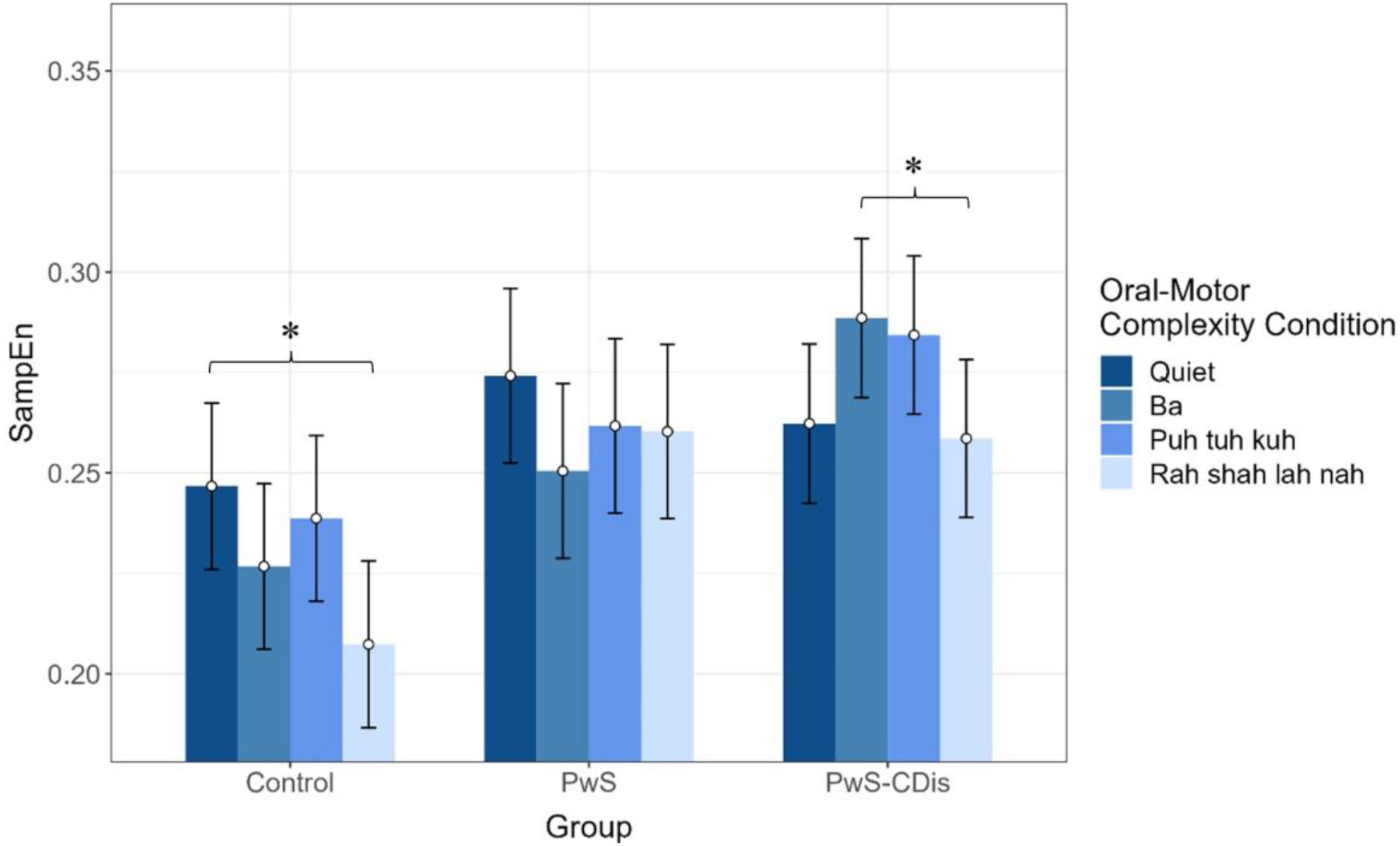
COP SampEn by group for each oral-motor complexity condition. Error bars represent the standard error of the mean (SEM). Asterisks (*) denote statistical significance (*p* < .05) between the “ba” condition and the “rah shah lah nah” in the PwS-CDis group and between quiet stance and the “rah shah lah nah” condition in the Control group.

## DISCUSSION

This study evaluated COP patterns in individuals with chronic stroke, both with and without communication disorders, during verbal utterance tasks of varying oral-motor complexity. Our hypothesis was partly supported: Individuals post-stroke with communication disorders exhibited differences in postural control dynamics across oral-motor complexity conditions compared to PwS without communication disorders and to a nondisabled control group. These differences were revealed primarily through nonlinear time-dependent metrics, indicating variations in the underlying control and organization of the postural control system as indicated by mathematical stability (MAXLINE), structural variation in behavioral modes (RQAEn), and regularity (SampEn). However, these postural control differences did not become more pronounced over conditions employing a linear progression in oral-motor complexity but instead varied depending on the group and the specific oral-motor task, pointing to a complex relation between speech-motor tasks and postural control. Notably, the PwS-CDis group demonstrated a distinct pattern of temporal dynamics across oral-motor complexity conditions that was not observed in participants who did not have stroke-related communication disorders.

Our results indicate that the PwS-CDis group consistently exhibited differences in the temporal dynamics of postural control between the “ba” condition (the lowest level of oral-motor complexity) and the “rah shah lah nah” condition (the highest level of oral-motor complexity). Specifically, the PwS-CDis group demonstrated increased mathematical stability (higher MAXLINE), greater structural regularity in behavioral modes (lower RQAEn), and a decrease in regularity (Vaillancourt & Newell, 2002) of postural sway patterns (higher SampEn) during the “ba” condition. With the exception of higher SampEn (indicating decreased regularity), the observed patterns are largely consistent with common postural control findings in neurological disability (e.g., Parkinson’s disease, multiple sclerosis) which are indicative of increased motor rigidity and reduced postural stability (Negahban et al., 2013; Perlmutter et al., 2010; Schmit et al., 2006; Schwab, Pinto, et al., 2021). This combination of findings suggests a more complex dynamic, possibly indicating that each postural control measure has an optimal value, and these results could reflect departures from that optimum (Harbourne & Stergiou, 2009; König et al., 2016; Slifkin & Newell, 1998; Stergiou & Decker, 2011; Stergiou et al., 2006), albeit in different directions for the different measures. In this context, when participants with stroke-related communication disorders performed the least complex oral-motor task, they concurrently exhibited COP patterns that were more mathematically stable and more consistent with regard to the number of behavioral modes exhibited, yet that were less regularly patterned over time. The PwS-CDis group also demonstrated higher mathematical stability compared to the Control group during both the “ba” condition and quiet stance, possibly indicating reduced flexibility and adaptability of movement in the PwS-CDis group during these conditions compared to nondisabled controls. These findings highlight the possible impact of oral-motor complexity on the underlying adaptability and organization of the postural control system in PwS with communication disorders.

There are a variety of potential reasons why PwS-CDis exhibited differences in temporal dynamics of postural control on the least complex speech task (“ba”) compared to the more complex speech tasks and compared to the Controls. One reason may be a potential respiratory interaction that occurred with each type of syllable complexity. There is a well-known coupling of breathing and speech, such that speakers take breaths at junctures of syntactic boundaries (such as between grammatical phrases) (Hird et al., 2002). For speakers in our study, respiration was not measured, but it was noted that speakers tended to treat the different multisyllable sequences “puh tuh kuh” and “rah shah lah nah” as if they were one word, where pauses (counted as breaths) occurred after and not within the pseudoword (i.e. “puh tuh kuh//puh tuh kuh” was evident and not “puh//tuh kuh”—where “//” marks a breath). It may be that the relatively simple oral-motor complexity of repeating “ba” was made more challenging for PwS-CDis—who have known limitations in speech and language—because of its lack of wordlike/grammarlike structure that would engage predictable speech respiratory behavior and the subsequent respiratory-postural coupling. Thus, the challenge of “ba” for people with post-stroke communication disorders may further manifest at the level of postural control by rigidly controlling postural dynamics. The results of our study highlight an important direction for future research to identify possible limitations in respiratory-postural coupling specifically in PwS with communication disorders. Whether the alterations in postural control observed in our study resulted from oral-motor task complexity alone, respiratory patterns, or their interaction, it is crucial to note that the changes in postural control emerged within the context of the specific oral-motor task complexity (“ba”), and it suggests a link between pathology, postural control, and speech motor tasks in individuals with post-stroke communication disorders, which was the focus of this specific study.

Interestingly, PwS-CDis demonstrated distinct differences in postural control temporal dynamics during the least and most complex speech tasks—but not during the task of medium complexity (“puh tuh kuh”). Task familiarity could potentially play a role in the performance on the medium complexity task, as the “puh tuh kuh” syllabic sequence is an assessment and treatment task (a diadochokinetic or DDK task) administered by speech-language pathologists to assess rate and regularity of syllable production (Kent, 2015). Therefore, PwS-CDis may have encountered this sequence in treatment for their communication disorder. However, the practice effects for DDK tasks are uneven, such that multiple repetitions of the task do not improve performance accuracy in older adults, though they do in young adults (Ben David & Icht, 2018). Therefore, traditional DDK sequences like “puh tuh kuh” may be less useful during clinical collaboration between physical therapists and speech-language pathologists as a dual task. Instead, a more complex sequence (such as “rah shah lah nah”) or the simple “ba” repetition task may present the appropriate level of challenge for a PwS-CDis.

In this study, the PwS-CDis group did not demonstrate significant differences in SD of COP, when compared to age-matched, nondisabled Controls. Additionally, increased SD of COP was found in the PwS group in the “ba” and “rah sha lah nah” conditions compared to the Control group; a difference notably absent in the PwS-CDis group. However, the differences found in the temporal structure and unfolding of postural control patterns in the PwS-CDis group (particularly MAXLINE) suggest an underlying difference in motor adaptability and flexibility during the performance of different motor speech tasks of varying complexity. This may provide important insights and clinical considerations for understanding the known alterations in balance, postural control, and fall risk among PwS with stroke-related communication disorders (de Haart et al., 2004; Dickstein et al., 2004; Schwab et al., 2023; Sullivan & Harding, 2019; Sullivan et al., 2021; Tasseel-Ponche et al., 2015). Overall, these findings underscore and further support the value of incorporating nonlinear time-series metrics like RQA and SampEn (e.g., Harbourne & Stergiou, 2009; Riley, 2001; Schmit et al., 2006) into studies investigating postural control post-stroke. Conducting nonlinear time-series analyses in the present study facilitated the curation of a more comprehensive picture of how oral-motor complexity may manifest in the temporal patterns of postural sway post-stroke.

The results of this study should be considered with respect to its limitations. This study categorized PwS with and without a history of concomitant speech disorders, without differentiating between mild, moderate, and severe categories or specifying the type of speech disorder (e.g., dysarthria vs. aphasia) or the location/severity of brain lesion. An interesting direction for future research may be to further stratify individuals with stroke-related communication disorders based on those subgroups to better understand how varying degrees and types of speech impairments/disorders might differentially impact postural control during speech tasks with different oral-motor complexities. Second, we did not objectively measure respiration, which could have helped clarify whether altered respiratory patterns across oral-motor complexity task levels may have impacted the results. Third, we did not control for the rate of utterances in different conditions, which precluded determination of whether utterance rate could have impacted our results. Lastly, our sample size was relatively small, which may affect the generalizability of results; null findings should be interpreted with caution.

## Conclusions

PwS with concomitant communication disorders demonstrated distinct temporal dynamics of postural sway during verbal utterance tasks of varying oral-motor complexity. The same changes across speech tasks of varying oral-motor complexity were not evident in PwS without communication disorders or nondisabled control participants. These findings highlight the potential impact of oral-motor complexity on the underlying adaptability and organization of the motor control system for people post-stroke with communication disorders, who exhibit a higher fall rate than individuals post-stroke who do not have communication disorders. Ultimately, this work contributes to the broader understanding of post-stroke rehabilitation, emphasizing the need for tailored approaches that address the multifaceted challenges faced by individuals with communication disorders, as distinct patterns of temporal dynamics in the PwS-CDis group suggests a possible link between pathology, postural control, and speech motor tasks after stroke.

## Data Availability

All data produced are available online at https://osf.io/n4acx/

## ACKNOWLEDGEMENTS

We sincerely thank the participants of this study for their time and willingness to participate.

## Footnotes

1 In written English, the number of letters and sounds may not exactly correspond. While “sh” is written with two letters, it is phonetically one sound.

